# Differential sensitivity to hypoxia enables shape-based classification of sickle cell disease and trait blood samples

**DOI:** 10.1101/2020.10.28.20221358

**Authors:** Claudy D’Costa, Oshin Sharma, Riddha Manna, Minakshi Singh, Samrat, Srushti Singh, Anish Mahto, Pratiksha Govil, Sampath Satti, Ninad Mehendale, Yazdi Italia, Debjani Paul

**Affiliations:** Department of Biosciences and Bioengineering, Indian Institute of Technology Bombay, Powai, Mumbai 400076, Maharashtra, India; MedPrime Technologies Pvt. Ltd., Casa Piedade Co-operative Housing Society, Charai, Thane – 400601, Maharashtra, India; Valsad Raktdan Kendra, Valsad - 396001, Maharashtra, India; Associate faculty, Wadhwani Research Centre for Bioengineering, Indian Institute of Technology Bombay, Powai, Mumbai 400076, Maharashtra, India

**Keywords:** sickle cell anemia, hemoglobin polymerization, chemically-induced hypoxia, microfluidics, smartphone microscopy, RBC shape, shape-based classifier

## Abstract

Differentiating between homozygous (disease) and heterozygous (trait) sickle cell patients is the key to ensuring appropriate long-term disease management. Well-equipped labs needed to perform confirmatory diagnostic tests are not available in endemic areas of most low- and medium-income countries. As a consequence of hemoglobin polymerization, red blood cells (RBCs) become sickle shaped and stiff under hypoxic conditions in sickle cell anemia patients. A simple test such as microscopy, using RBC shape as a biophysical marker, cannot conclusively differentiate between homozygous (disease) and heterozygous (trait) sickle blood. Here, we establish a new paradigm of microscopic diagnosis of sickle cell disease by exploiting differential polymerization of hemoglobin in disease and trait RBCs under controlled, chemically-induced hypoxia in a microfluidic chip. We use a portable smartphone microscope to compare the RBC shape distributions in blood treated with high and low concentrations of the hypoxia-inducing agent to correctly identify 35 blood samples as healthy, sickle cell disease or trait. Finally, we demonstrate our test in remote field locations to enable fast and confirmed diagnosis of sickle cell anemia in resource-limited areas.

## Introduction

Sickle cell anemia is a genetic disorder caused by a glutamine to valine mutation in the β-globin gene^1^. It results in partial or complete replacement of normal adult hemoglobin (HbA) with mutated sickle hemoglobin (HbS). HbS polymerizes under hypoxic conditions and forms rigid and misshapen red blood cells (RBCs). The loss of deformability of RBCs leads to frequent vaso-occlusive crisis, joint pain, spleen damage, increased susceptibility to infection and anemia. While there is no cure available for sickle cell anemia, early diagnosis can prevent child mortality and improve the quality of life of the affected individuals.

Sickle cell disease is widely prevalent in several parts of the world including western Africa, Latin America, the Arab peninsula, and India^2^. More than 500 children die of sickle cell anemia every day due to lack of comprehensive newborn screening programs in low- and medium-income countries (LMIC). Distinguishing between sickle cell disease (homozygous) and trait (heterozygous) individuals is crucial to ensure appropriate clinical and non-clinical interventions. The current two-step diagnosis protocol involves a solubility test to screen for HbS-positive individuals, followed by hemoglobin electrophoresis or high-performance liquid chromatography (HPLC) to distinguish between homozygous and heterozygous individuals^3^. The laboratory infrastructure and trained personnel required for these tests are not always available in LMICs to sustain countrywide screening programs^4^.

Preparation of blood smear slides and their microscopic examination are diagnostic techniques that are relatively easier to administer in LMICs. Despite the widespread use of microscopy to study sickle blood in the early days^2–5^, it is no longer a technique preferred by the clinicians. Sickle blood has only 5% - 25% irreversibly sickled cells (ISCs)^6^, requiring extensive scanning of each smear slide by a technician. While use of a chemical oxygen scavenger, such as sodium dithionate or sodium metabisulphite, allows more RBCs to sickle *in vitro*, the process can take as long as 24 hours for trait blood samples. Most importantly, it is not possible to conclusively distinguish between sickle cell disease and trait blood by simple visual inspection of RBC shapes.

Several research groups have explored image processing or machine learning techniques to study RBC morphologies^7–18^. These techniques have focused on classification of individual RBC shapes into normal (biconcave), sickle or abnormal (i.e. having any shape other than biconcave or sickle). None of these reports have used RBC shape as a sole biophysical marker to classify sickle blood samples into disease or trait. Recently, Javidi and others classified blood samples into healthy and sickle based on membrane fluctuation analysis of individual RBCs^19^. De Haan *et al* developed a mobile phone microscope to image blood smears, followed by automated identification of ISCs in these images^20^. As a shortcoming of their method, the authors stated that blood smears cannot be used to identify sickle cell genotypes, i.e. to discriminate between trait and disease blood samples. There is currently no test that uses RBC shape as a sole biophysical marker to classify sickle blood samples into disease or trait.

Here, we combine microfluidics, smartphone microscopy and image processing techniques (**figure 1**) to develop a method that accurately distinguishes between disease and trait sickle blood based on the differential shape changes of RBCs when treated with two different concentrations of an oxygen scavenger, sodium metabisulphite. As shown in **figure 1A**, the method consists of mixing a drop of blood with a specific concentration of sodium metabisulphite, loading it into a microfluidic chip and imaging the sickled RBCs in real time using a smartphone microscope. The roundness distributions of RBCs after 30 min are used to classify the blood sample as healthy, disease or trait. The use of a microfluidic chip (**figure 1B**) as an imaging chamber instead of a smear slide enables fast sickling and tracking of RBC shapes in real time. The chip is made of glass to facilitate brightfield imaging of unstained RBCs using a mobile phone microscope and to maintain hypoxic conditions inside the chamber for the duration of the experiment. We optimized the height of the chamber to be 30 μm to avoid stacking of RBCs. We also designed and fabricated a portable smartphone microscope (**figure 1C**) to conduct our experiments outside the laboratory at sickle cell screening camps.

**Figure 1.**
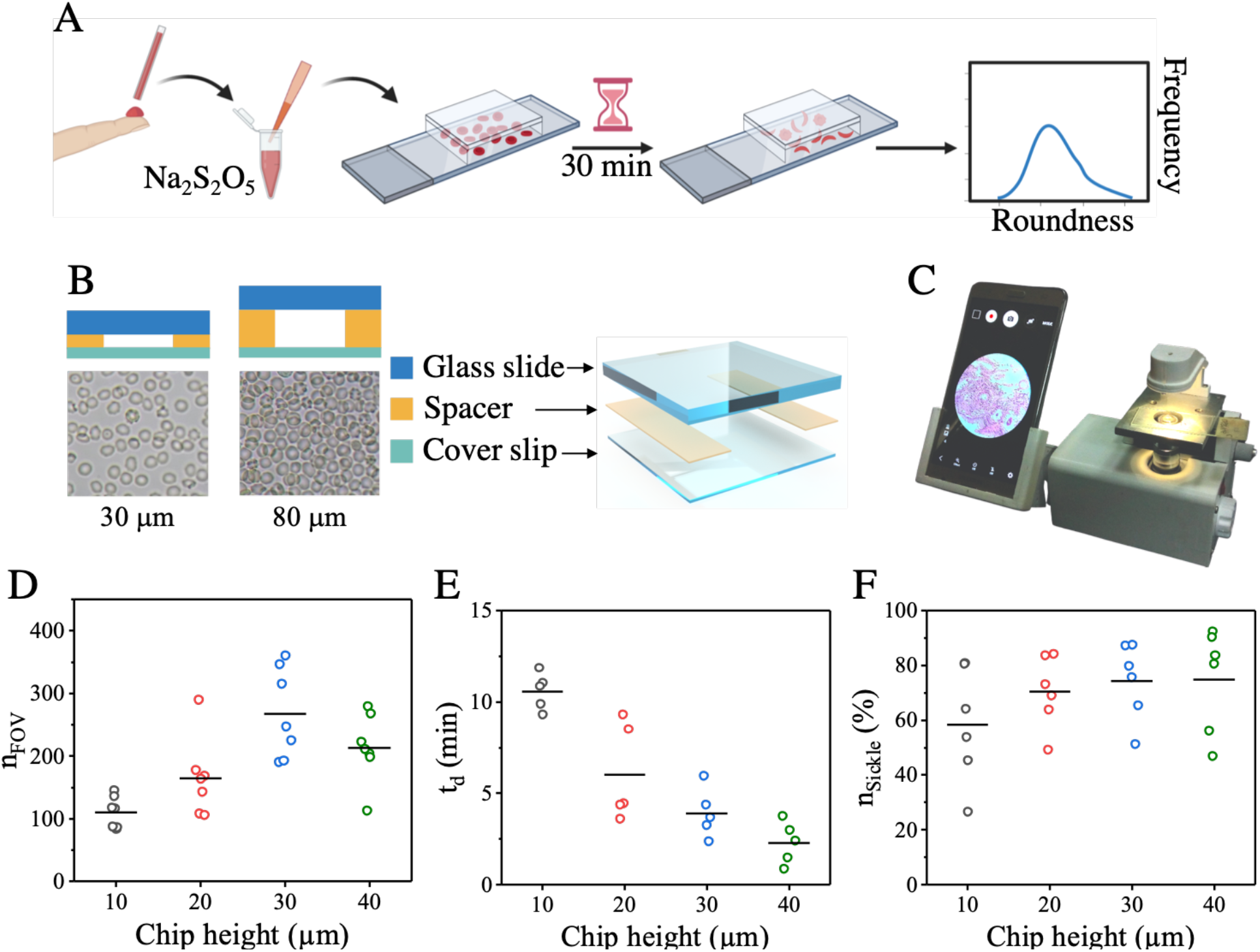
An assay to induce controlled sickling of RBCs inside a microfluidic chip, followed by imaging and analysis of RBC shapes. (A) A drop of blood is added to an oxygen scavenger (Na_2_S_2_O_5_), introduced into a microfluidic chamber and sealed. We image the sample after 30 min and analyze the RBC shapes to obtain a characteristic roundness distribution. (B) Exploded view of the microfluidic chamber. The thickness of the spacer determines the height of the chamber. RBCs inside a 30 μm high chamber are oriented flat, while they are stacked and randomly oriented when the height is 80 μm. (C) A photo of the portable smartphone microscope designed and fabricated by us. (D) Number of healthy RBCs in the field of view (n_FOV_) inside imaging chambers of different heights (N =7). (E) The time taken (t_d_) for the first RBC in the field of view to start sickling inside chips of different heights (N =6). (F) The percentage of sickle cells (n_**sickle**_) at t = 30 min inside chips of different heights (N = 6). The horizontal lines indicate the mean values in panels D-F.

## Methods

### Blood samples

All studies using human blood samples were approved by the Institute Ethics Committee (IEC), IIT Bombay, with approval numbers IITB-IEC/2016/016, IITB-IEC/2017/020 and IITB-IEC/2018/042. Leftover de-identified blood samples were collected from adults and older children with a mix of genders during sickle cell screening camps organized by Shirin and Jamshed Guzder Regional Blood Centre (Valsad, Gujarat) and Dayanand Hospital (Talasari, Maharashtra). Written informed consent to use leftover blood samples was taken from all participants. The ages of the participants ranged from 10 to 60 years. We excluded those individuals who (i) were under treatment with folic acid or hydroxyurea, or (ii) received blood transfusion within three months prior to blood collection. Blood samples from self-reported healthy volunteers from IIT Bombay and Dayanand Hospital were used as healthy samples during development and validation of the classifier. All known sickle blood samples (D = 23 and T = 92) were first screened by the hospital personnel using a standard solubility test. Their hemoglobin profiles were then confirmed using HPLC to conclusively identify them as disease or trait.

### Experiment protocol

Blood samples collected in K3/EDTA vacutainer tubes were stored at 4°C. All tests were performed either at IIT Bombay or in field locations within 48 h of blood collection. Sodium metabisulphite solutions were freshly prepared in cell culture media (RPMI-1640) before each experiment and discarded after 3 h. A drop of blood was added to 0.1% and/or 0.3% sodium metabisulphite to dilute it by 20X, irrespective of hematocrit value. The average pH of blood mixed with 0.1% and 0.3% sodium metabisulphite solution was 6.05 ± 0.2 and 5.83 ± 0.1 respectively. A 10 µl volume of 20X diluted blood mixed with an appropriate concentration of sodium metabisulphite was introduced into the imaging chamber and the chip was sealed with quick-drying transparent nail lacquer. This step was repeated if air bubbles got trapped in the sealed chip or significant crenation occurred in RBCs immediately after loading. The chip was placed on the sample stage of a smartphone microscope and images of RBCs were captured at 0 min and 30 min. We also captured real-time videos of sickling for 30 min at 30 fps to understand the sickling kinetics. While the ambient temperature in the lab ranged from 29°C to 31°C, the temperature during field testing varied between 25°C and 32°C. High performance liquid chromatography (HPLC) was performed at Shirin and Jamshed Guzder Regional Blood Centre on all sickle blood samples to obtain their hemoglobin profiles (HbA, HBA_2_, HbS and HbF).

### Mobile phone microscope

We developed a battery-operated inverted transmission microscope to capture images of unstained RBCs in the field. The microscope has a 1W white LED, fitted with a collimating lens, as the illumination source and a single 40X (0.65 NA) air objective. The image is focussed on the mobile phone camera by vertically moving the sample stage. We used Xiaomi Mi3 or Samsung A10 mobile phones to capture images of RBCs with a minimum resolution of 2368 pixels *×* 4208 pixels. These phone models were chosen to balance performance, cost and availability in developing countries. We developed one microscope model with a fixed phone holder and another where the phone holder can be tilted by any angle for viewing comfort. The sample stage and the base of the microscope are made of aluminium for increased stability, while the remaining parts, including the outer body, are 3D-printed using PLA. Exploded images of the mechanical and optical parts of the microscope can be seen in **figure S1** in supplementary information.

### Image analysis and classification

Brightfield images of unstained RBCs were analyzed by ImageJ following a process flow shown in **figure S2** in the supporting information^21^. We measured roundness and solidity of each RBC. RBCs with solidity <0.8 were excluded from analysis as these corresponded to partially focused RBCs. Roundness values of remaining ∼ 150 -200 RBCs in the field of view were binned into 10 groups with 0.1 width to obtain the roundness distribution. Each distribution was normalized by dividing by the total number of RBCs included in the analysis. We used R programming language^22^ to calculate connectivity and Dunn index for all 16 P_1_ and P_2_ combinations. The connectivity was calculated with 10 nearest neighbors. By applying a support vector machine (SVM) model with a linear kernel and a cost of 10 on the raw data, we obtained a classifier given by the equation: y = –1.527x + 0.633.

## Results and discussion

### The optimized height the microfluidic imaging chamber is 30 µm

We fabricated our imaging chambers using glass due to its non-permeability to oxygen and good optical properties. We obtained different chamber heights (10 µm, 20 µm, 30 µm, 40 µm, 80 µm and 100 µm) using double-sided adhesive films of specific thicknesses as spacers. An increase in microfluidic confinement increased crenation of healthy RBCs. A chamber height of 80 µm or above reduced crenation and promoted sickling, but led to stacking of RBCs. Therefore, we focussed on chips of intermediate heights, e.g. 10, 20, 30 and 40 µm.

As shown in **figure 1D**, we counted the number of RBCs in the field of view (*n*_*FOV*_) inside chambers of different heights. Relatively fewer RBCs were present inside 10 µm (110 ± 10; mean ± SEM) and 20 µm (165 ± 23) high chambers. In contrast, the image processing algorithm detected more than 200 RBCs inside chambers with 30 µm (268 ± 27) and 40 µm (213 ± 21) heights. The number of RBCs detected inside the chamber increases with an increase in the chamber height. As there are more densely packed RBCs inside the 40 µm high chamber, these are eliminated as clusters by the image processing algorithm. This results in a decrease in the number of RBCs counted by the algorithm inside the 40 µm high chamber.

As shown in **figure 1E**, we treated six sickle cell disease blood samples with 0.1% sodium metabisulphite. From real time videos of sickling, we measured the time (*t*_*d*_) it takes for the first RBC in the field of view to sickle inside imaging chambers of different heights. A detailed discussion on the delay time is given elsewhere in the paper. The values of *t*_*d*_ are 10.6 ± 0.5 min (mean ± SEM), 6.0 ± 1.2 min, 3.9 ± 0.6 min and 2.3 ± 0.5 min for chip heights of 10, 20, 30 and 40 µm respectively.

We also measured the percentage of sickled RBCs (*n*_***sickle***_) at t = 30 min inside these chambers. As shown in **figure 1F**, the mean value of *n*_***sickle***_ ranged from 58% to 75% with a large variability in the data. We found that RBCs inside a 30 µm chamber do not crenate much and sickle relatively fast (*t*_*d*_ = 3.9 ± 0.6 min). On an average, there are > 250 RBCs in the field of view. Therefore, all further experiments in this study were conducted inside 30 µm chambers.

### Difference in polymerization of sickle hemoglobin in disease and trait blood leads to different RBC shapes

I. J. Sherman first connected the shapes of sickled RBCs to the rate of oxygen removal by noting that slow deoxygenation resulted in RBCs with sickle shapes, while sudden deoxygenation made them granular. He reported that disease blood had more sickled RBCs in it compared to trait blood^6^. A relation between RBC shapes and polymerization kinetics of sickle hemoglobin (HbS) was later proposed by Eaton and Hofrichter. Based on extensive *in vitro* studies on HbS, they predicted that sickle shapes of RBCs are likely to result from the growth of a single polymer domain during slow polymerization. Presence of multiple small polymer domains with shorter fibers should lead to holly leaf cell shapes. When there are many more randomly oriented and very short fibers, RBCs become granular^23^. Direct experimental evidence of this prediction using linear dichroism microscopy was provided by Mickols *et al*^24^ and by Corbett *et al*^25^. They measured the distribution and orientation of aligned hemoglobin polymer domains inside RBCs and related it to different cell shapes.

**Figure 2** schematically illustrates different HbS polymerization mechanisms in disease and trait blood and the associated RBC shapes. The total hemoglobin in homozygous sickle blood consists of 95% - 98% HbS, 2% - 3% HbA_2_ (another variant of normal hemoglobin) and 2% HbF (fetal hemoglobin). Heterozygous sickle cell blood contains 35% - 45% HbS, 50% - 65% HbA, 2% - 3% HbA_2_ and ∼ 2% HbF^26^. As shown in **figure 2A**, blood from healthy individuals has HbAA homodimers (*α*_2_*β*_2_), sickle cell disease blood has HbSS homodimers (*α*_2_*β*^S^_2_), and trait blood contains a mix of HbAA homodimers (*α*_2_*β*_2_), HbSS homodimers (*α*_2_*β*^S^_2_) and HbAS heterodimers (*α*_2_*ββ*^S^).

**Figure 2.**
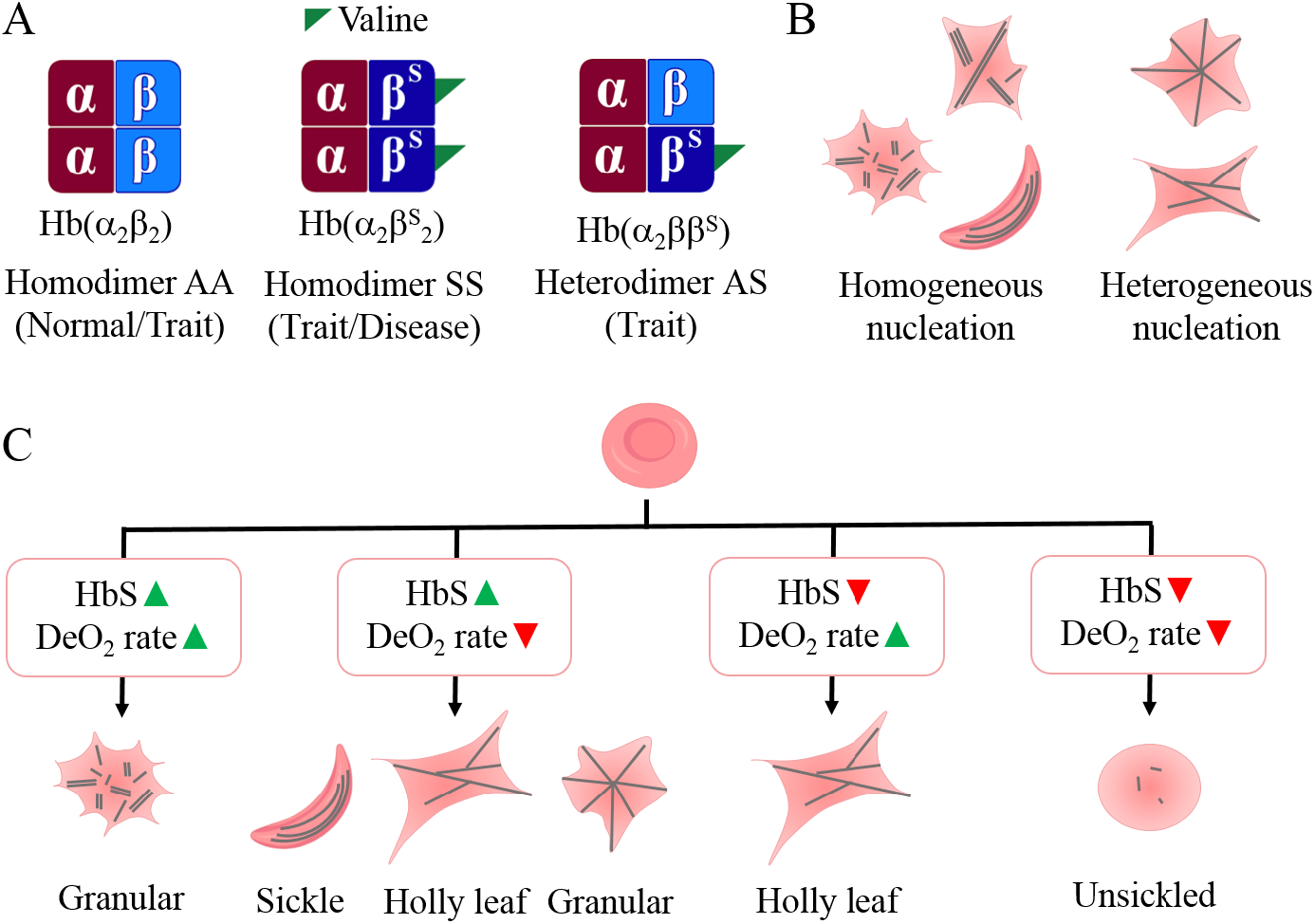
Schematic diagram relating hemoglobin polymerization mechanism to the shapes of sickled RBCs. (A) Hemoglobin in healthy blood exists as homodimer AA (α_2_β_2_). This homodimer is also found in trait blood. Sickle hemoglobin can either form a homodimer SS (α_2_β^S^_2_), found in both trait and disease blood, or a heterodimer AS (α_2_ ββ^S^) with normal hemoglobin, found only in trait blood. (B) During hemoglobin polymerization, homogeneous nucleation leads to one or more non-branched polymer chains, while heterogeneous nucleation leads to branched polymers. The kind of hemoglobin polymerization that takes place inside an RBC affects its shape. (C) A pictorial summary of how shapes of sickled RBCs depend on HbS concentration and deoxygenation rate. Green ‘up triangle’ symbol indicates a high HbS concentration or fast deoxygenation. Red ‘down triangle’ symbol indicates low HbS concentration or slow deoxygenation.

According to the double nucleation model proposed by Ferrone *et al*, hemoglobin polymerization can proceed by homogenous or heterogeneous nucleation (**figure 2B**)^27^. Homogeneous nucleation involves growth of independent polymer chains from HbS molecules in solution, whereas heterogeneous nucleation involves formation of branches on already existing polymers. Even though homogeneous nucleation is thermodynamically less favorable, Ferrone and others suggested that it can be sustained in samples with high initial HbS concentration for a longer duration before heterogeneous nucleation takes over. Since sickle cell disease blood has high HbS concentration, it follows that these samples are likely to sustain homogeneous nucleation for longer. The authors further observed that fast deoxygenation leads to more homogeneous nucleation sites and formation of randomly oriented polymer chains. Hence, we expect that faster deoxygenation in disease samples would lead to more granular RBCs, and very few, if any, sickle-shaped RBCs. Slow deoxygenation in these samples would lead to sickle RBCs, resulting from sustained homogeneous nucleation, as well as some holly leaf and granular RBCs formed due to heterogeneous nucleation.

In addition to HbS concentration and the rate of deoxygenation, HbS polymerization in trait samples is also affected by the presence of HbAS heterodimers which can constitute as much as ∼ 49% mole fraction of the total hemoglobin^28^. HbS concentration in trait samples is too low to sustain homogeneous nucleation. Moreover, HbAS heterodimers have a lower probability to take part in polymerization compared to HbSS homodimers^29^. Under slow deoxygenation, due to a lack of nucleation sites, RBC morphology in trait blood appears to be unchanged. When deoxygenation is rapid, trait RBCs deform into primarily holly leaf shapes resulting from heterogeneous nucleation. **Figure 2C** summarizes the combined effect of HbS concentration and rate of deoxygenation on RBC shapes. Our shape-based classification method to identify unknown blood samples makes use of the variation in RBC shapes in disease and trait samples in response to slow and fast deoxygenation.

### Specific concentrations of sodium metabisulphite can lead to slow and fast deoxygenation

We optimized the concentration (w/v) of sodium metabisulphite with the goal of identifying two specific concentrations at which disease and trait samples are likely to undergo slow and fast deoxygenation resulting in different RBC shapes. **Figure 3A** shows a plot of the dissolved oxygen content as a function of time for different concentrations of sodium metabisulphite. For the entire range of concentrations studied, the oxygen content decreased to <5% within 30 min, which is considered to be the physiological level of deoxygenation in blood^30^.

**Figure 3.**
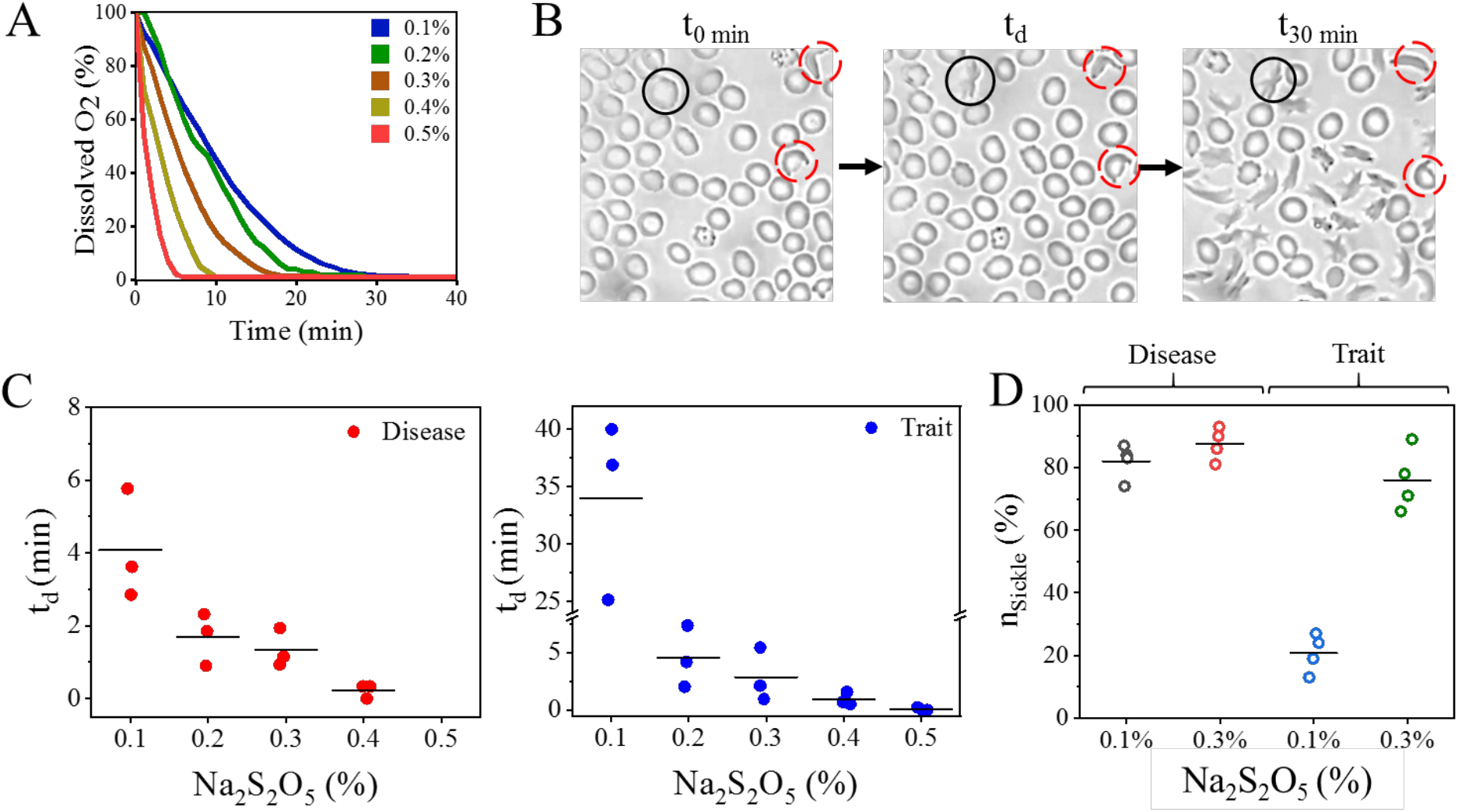
Optimization of sodium metabisulphite concentration to induce slow and fast deoxygenation in disease and trait blood samples. (A) Decrease in the dissolved oxygen in cell culture media (RPMI-1640) with time for different concentrations of sodium metabisulphite. (B) Schematic representation of the time (t_d_) taken by the first unsickled RBC (enclosed by the black circle) in the field of view to sickle. Irreversibly sickled RBCs are encircled in dashed red. (C) t_d_ as a function of sodium metabisulphite concentration in three distinct disease and three distinct trait samples, represented as mean ± SEM (N = 3). (D) Number of sickled RBCs in disease (N = 4) and trait (N = 4) samples treated with 0.1% and 0.3% sodium metabisulphite respectively. The horizontal lines indicate the mean values in all the plots in (C) and (D).

We then mixed 0.1% to 0.5% sodium metabisulphite solutions with disease and trait blood samples (**table S1** and **figure S3** in supporting information). We recorded real time sickling videos in our smartphone microscope, and from the extracted frames, determined the instant (*t*_*d*_) at which the first unsickled RBC in the field of view starts sickling. **Figure 3B** shows how *t*_*d*_ is measured for the RBC indicated by the solid black circle, with the first frame in the video taken as the timepoint *t* = 0 min. RBCs inside the dashed red circles are already sickled, and therefore, are not considered for this purpose.

We then used *t*_*d*_ as a parameter to compare the sickling behavior of disease and trait RBCs treated with different sodium metabisulphite concentrations. **Figure 3C** shows how *t*_*d*_ varies with sodium metabisulphite concentration for three disease and three trait samples. For disease samples treated with 0.1%, 0.2% and 0.3% concentrations, *t*_*d*_ is 4.1 min ± 0.9 min (mean ± SEM), 1.7 ± 0.4 min and 1.3 ± 0.3 min respectively. The corresponding values for trait samples are 33.4 ± 4.8 min, 4.5 ± 1.5 min and 2.8 ± 1.3 min respectively. Since sickling occurs almost instantaneously at 0.4% and 0.5% concentrations, making it difficult to accurately measure *t*_*d*_, these two concentrations were not considered further.

While 82 ±3 % (mean ± SEM) of RBCs in disease blood sickle within 30 min when treated with 0.1 % concentration, only 21 ± 3% of RBCs in trait blood sickle at this concentration (**figure 3D**). In contrast, the average number of sickled RBCs in disease and trait blood samples treated with 0.3% sodium metabisulphite for 30 min are 88 ± 3 % and 76 ± 5% respectively.

Therefore, based on the differential sickling response of RBCs in our experiments, we chose 0.1% and 0.3% as the two sodium metabisulphite concentrations to induce slow and fast deoxygenation respectively in sickle blood samples. The experiments on quantifying the shape distribution of RBCs were performed with these two sodium metabisulphite concentrations.

### Deoxygenated healthy, disease and trait blood have characteristic roundness distributions

We treated three samples each of healthy, disease and trait blood with 0.1% and 0.3% sodium metabisulphite and imaged the RBCs after 30 min. Images of RBCs in the same blood samples, without addition of the oxygen scavenger, were used as negative controls. **Figure 4A** shows snippets of representative images from healthy, disease and trait blood samples, while the shapes of individual sickled RBCs are shown in **figure 4B**. The complete raw images from which the snippets are taken are shown in **figure S4** in the supporting information.

**Figure 4.**
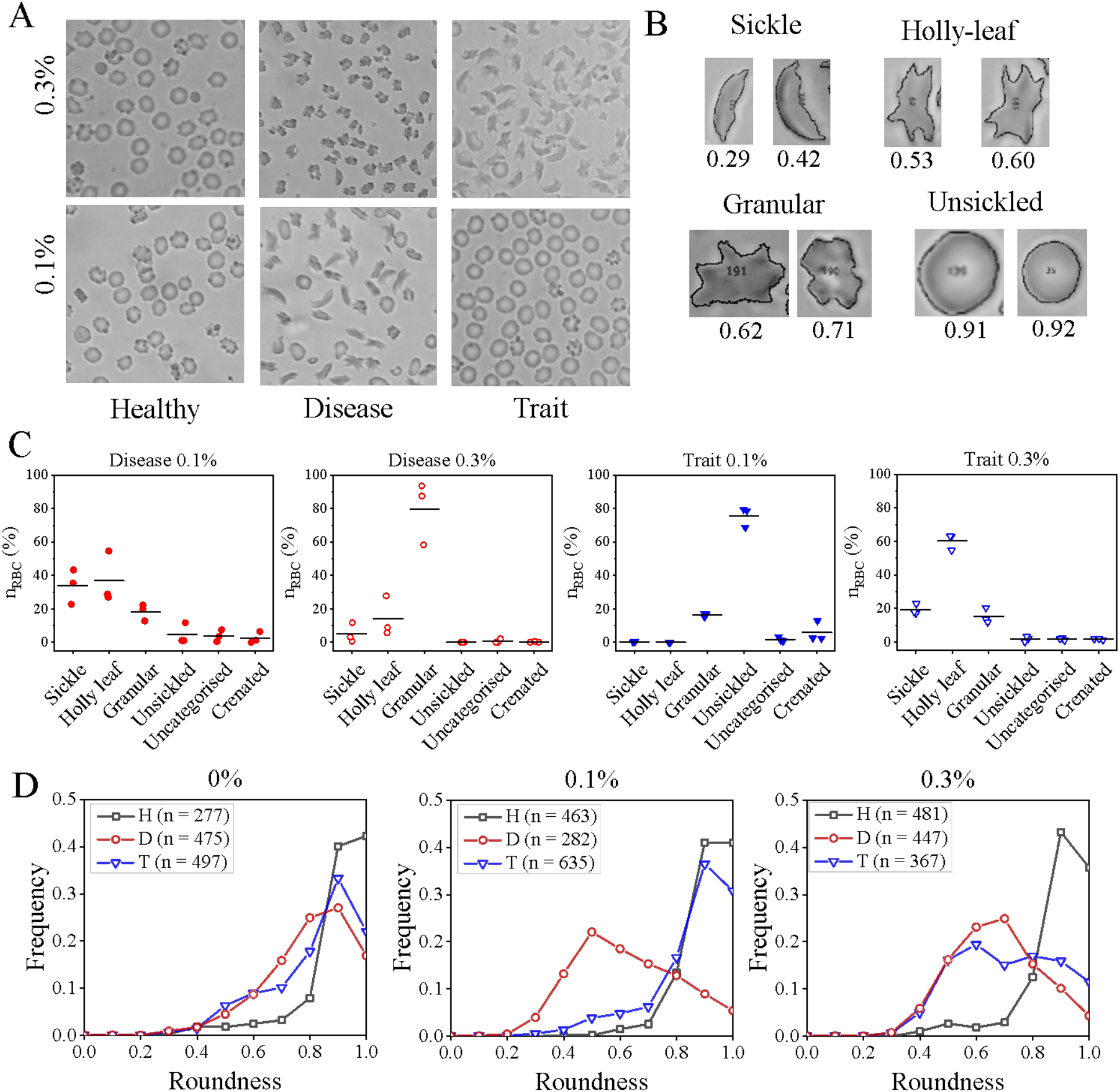
Roundness as a shape descriptor for quantifying RBC shapes in healthy and sickle blood. (A) Snippets of representative images of healthy, disease and trait samples treated with 0.1% and 0.3% sodium metabisulphite for 30 min are shown. Some crenated RBCs are seen in both the healthy samples and the trait blood sample treated with 0.1% sodium metabisulphite. (B) Sickle RBCs under hypoxia have a range of shapes which can be characterized by a roundness value. (C) Distribution of different kinds of RBC shapes in deoxygenated disease (N = 3) and trait (N = 3) treated with 0.1% and 0.3% sodium metabisulphite. The horizontal lines indicate the mean values. (D) Roundness distributions of RBCs in healthy (H; black square), disease (D; red circle) and trait (T; blue triangle) samples treated with 0%, 0.1% and 0.3% sodium metabisulphite respectively are shown. Each distribution is generated by analysing three different blood samples (N =3) and then pooling the data to see the overall trend. Here, n indicates the total number of RBCs in three samples used to generate each roundness distribution plot. Each plot is normalized by dividing it by the number of RBCs (n).

An experienced user then manually annotated and counted the number of sickle-shaped, holly leaf-shaped, granular, and unsickled RBCs in each of these disease and trait images. **Figure S5** in supporting information shows a typical annotated image. Since we found that the presence of crenated RBCs leads to image artifacts, we also accounted for these RBCs in each sample. RBCs which could not be classified into any of these categories were marked as ‘uncategorised’. As shown in **figure 4C**, disease samples treated with 0.1% sodium metabisulphite have primarily a mix of sickle (34 ± 6%; mean ± SEM), holly leaf (37 ± 9%) and granular (18 ± 3%) RBCs. The same samples, when treated with 0.3% concentration, have primarily granular (80 ± 11%) and a few holly leaf (14 ± 7%) RBCs. Trait samples treated with 0.1% metabisulphite have (76 ± 4%) unsickled RBCs. These samples have (19 ± 2%) sickle, (60 ± 3%) holly leaf and (15 ± 3%) granular RBCs when treated with 0.3% metabisulphite concentration.

Wheeless *et al* identified ‘form factor’ (*FF*), given by equation (1), to be the best image analysis feature to distinguish between sickle RBC shapes after comparing 42 shape descriptors^31^. As shown in **figure S6** in the supporting information, ‘roundness’ (*R*), given by equation (2), is a better measure of RBC shapes in our images than form factor.

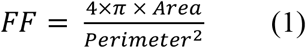

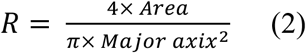

**Figure 4D** shows the roundness distribution plots of RBCs in healthy (indicated by black square), disease (red circle) and trait (blue triangle) blood, where each curve has data pooled from three samples. Here, *n* indicates the number of RBCs from these three samples used to generate each distribution plot. The left panel shows the control distributions in absence of the oxygen scavenger. The distribution corresponding to healthy blood peaks at 1.0 as healthy biconcave RBCs lying flat inside the imaging chamber appear circular in shape. The distributions for untreated trait and disease samples peak at 0.9 due to the presence of very few ISCs. The disease distribution has a broader peak compared to the trait distribution as disease samples have more ISCs than trait samples.

The middle panel shows the roundness distributions of blood samples treated with 0.1% sodium metabisulphite. Healthy RBCs show a peak at 0.9 – 1.0 as these do not sickle when treated with an oxygen scavenger. Due to a combination of slower deoxygenation and high HbS concentration, RBCs in disease samples have a mix of sickle, holly leaf and granular shapes, resulting in a broad distribution with a peak at 0.5. The shapes of most RBCs in trait blood samples treated with 0.1% sodium metabisulphite remain unchanged owing to the combined effect of fewer nucleation sites, HbAS heterodimers, and slow deoxygenation. As a result, the roundness distribution peaks at 0.9, similar to healthy RBCs.

The right panel shows roundness distributions of RBCs treated with 0.3% sodium metabisulphite. A combination of faster deoxygenation and higher HbS concentration in disease samples results in granular RBCs, which is reflected in a broad distribution with a peak at 0.7. On the other hand, owing to faster deoxygenation and lower HbS concentration, RBCs assume holly leaf shapes in trait samples. This leads to a somewhat bimodal roundness distribution with peaks at 0.6 and 0.8 respectively. As expected, the distribution for healthy RBCs has a peak at 0.9. The tails of the distributions for healthy RBCs at lower roundness values in all three panels result from certain analysis artifacts, as shown in **figure S7** in the supporting information.

As RBCs with holly leaf and granular shapes have similar roundness values, the roundness distributions of trait and disease samples treated with 0.3% sodium metabisulphite overlap with each other. Similarly, the peaks of roundness distributions corresponding to healthy and trait samples treated with 0.1% sodium metabisulphite overlap.

### A shape-based classifier distinguishes between healthy and sickle blood

We derived two secondary shape parameters *P*_1_ and *P*_2_ from each roundness distribution, using a process flow shown schematically in **figure 5A**. We identified two specific areas under each roundness distribution curve by placing two contiguous windows of equal width (left panel). We called these two areas parameter 1 (*P*_1_; violet) and parameter 2 (*P*_2_; green) respectively. Each roundness distribution is represented as a single point on a plot of *P*_2_ vs. *P*_1_. We then translated the pair of windows by increments of 0.1 over the entire range of roundness values, while also varying their widths from 0.2 to 0.5, and calculated all sixteen *P*_1_ - *P*_2_ combinations resulting from this operation (middle panel). Widths <0.2 were not considered as they would contain information about very few RBCs. We then ran a k-means algorithm on each of these 16 datasets to distinguish between heathy and sickle (e.g. disease and trait) clusters. The optimal *P*_1_ - *P*_2_ combination was identified using connectivity and Dunn index. Connectivity indicated how strongly two clusters were connected, while Dunn index indicated intra-cluster compactness and inter-cluster separation^32^. We finally plotted Dunn index vs. connectivity to identify the most robust combination with low connectivity and high Dunn index (right panel) lying in the top right quadrant of the plot.

**Figure 5.**
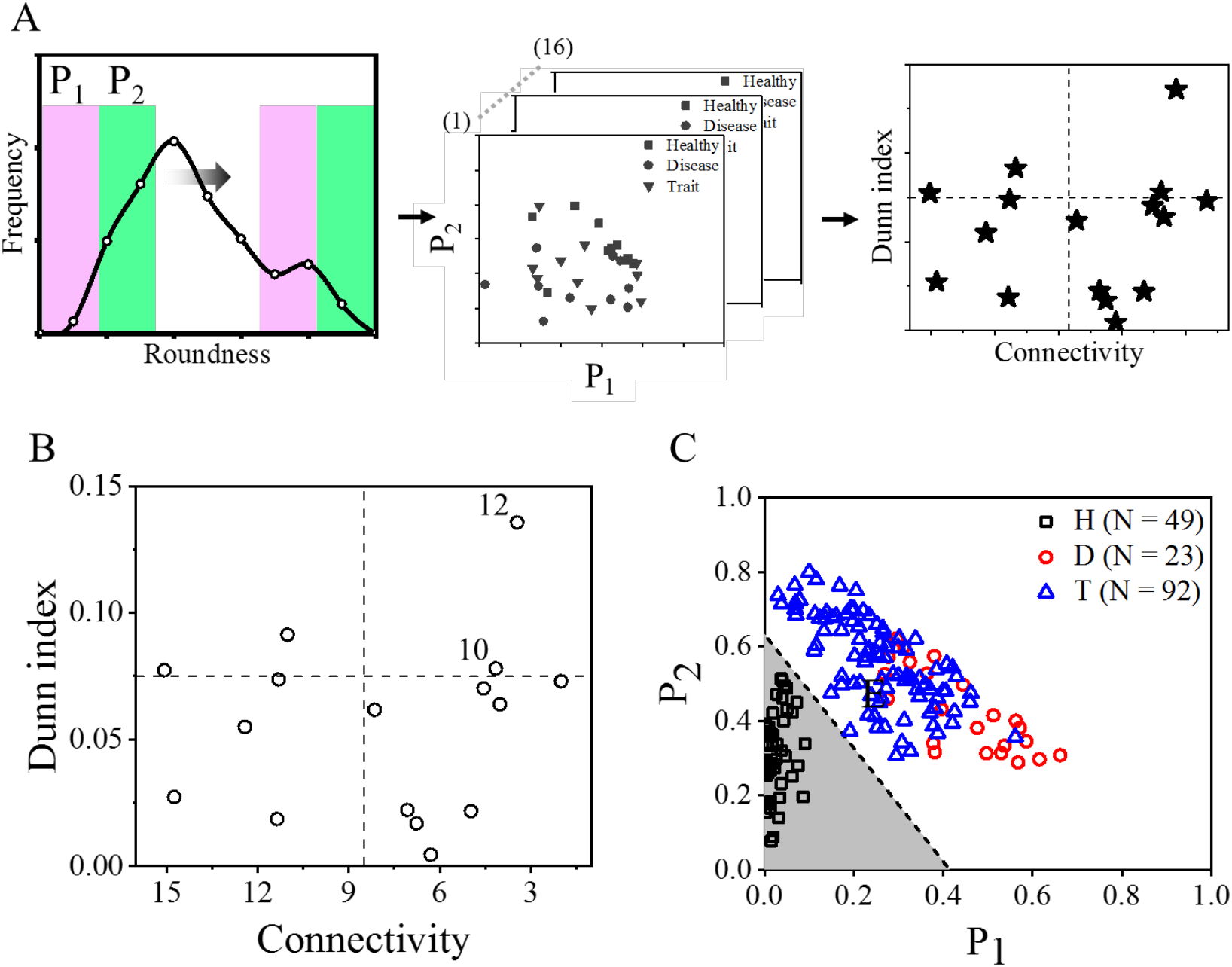
Derivation of two secondary shape parametersP_1_ and P_2,_ from the roundness distributions. (A) Schematic diagram describing the process flow to choose a robust combination of P_1_ and P_2_. (B) A plot of Dunn index vs connectivity for 16 unique combinations of P_1_ and P_2_ corresponding to 164 blood samples. The top-right quadrant of the plot indicates the most robust combinations (e.g. 10 and 12) with low connectivity and high Dunn index values. (C) Development of the classifier. P_2_ vs P_1_ data for 164 distinct blood samples with known haemoglobin profiles are plotted. Two distinct clusters are seen, one for the normal samples (HbA), and one for the trait and diseased samples (HbS). The classifier (dotted line) separates the parameter space into two regions indicated in grey and white.

**Figure 5B** shows a plot of Dunn index vs. connectivity for 16 *P*_1_ − *P*_2_ combinations generated from the roundness distributions of 164 blood samples (H = 49, D = 23 and T = 92) with known hemoglobin profiles. The values of the parameters used in this plot are given in **table S2** in the supporting information. For this experiment, we treated disease samples with 0.1% sodium metabisulphite as the roundness distributions of disease and trait samples are different at this concentration. Similarly, both trait and healthy samples were treated with 0.3% sodium metabisulphite to distinguish them from each other.

There are two points in the top right quadrant corresponding to the roundness ranges of 0.2 – 0.8 (combination #10; *P*_1_1 0.2 − 0.5; *P*_2_1 0.5 − 0.8) and 0.4 − 1.0 (combination #12, *P*_1_1 0.4 − 0.7; *P*_2_1 0.7 − 1.0) respectively that meet the selection criteria. Combination #10 includes information about sickle RBCs with 0.2<R<0.4 and excludes information about unsickled RBCs with R>0.8. Combination #12, with the highest Dunn index, lacks information about sickle RBCs with R<0.4, but includes information about unsickled RBCs with R>0.8. As combination #10 contains information about RBCs that are physiologically more relevant to our study, we preferred it over combination #12.

**Figure 5C** shows the *P*_2_ vs. *P*_1_ plot of 164 known samples corresponding to the first combination. We used a support vector machine (SVM) model on this dataset to develop a classifier shown by the dotted line that distinguished between healthy and sickle (e.g., both disease and trait) blood. A point that lies in the grey parameter space below the classifier should correspond to a blood sample with unsickled RBCs, while a point that lies in the white parameter space above the classifier should correspond to a blood sample with sickled RBCs.

### Distinguishing between disease and trait blood using our classifier

Based on our analysis, we proposed the work flow shown in **figure 6A** to classify an unknown blood sample. As demonstrated in the previous sections, it is not possible to unambiguously identify an unknown blood sample as healthy, disease or trait by treating it with just a single concentration of sodium metabisulphite. Therefore, we treat each unknown sample with both 0.1% and 0.3% sodium metabisulphite and plot the corresponding *P*_1_ and *P*_2_ values. As healthy RBCs do not sickle with either concentration, *P*_1_ and *P*_2_ are low, and the corresponding points (hollow and solid black squares) lie in the grey region below the classifier. In contrast, more than 80% RBCs in disease samples sickle when treated with either 0.1% or 0.3% sodium metabisulphite, resulting in high P_1_ and P_2_ values. Hence, both points (hollow and solid red circles) corresponding to disease samples lie in the white region above the classifier. As very few RBCs in trait samples sickle when treated with 0.1% sodium metabisulphite, the corresponding point (solid blue triangle) lies below the classifier. The same trait sample has high P_1_ and P_2_ values when treated with 0.3% sodium metabisulphite. Therefore, the corresponding point (hollow blue triangle) lies above the classifier.

**Figure 6.**
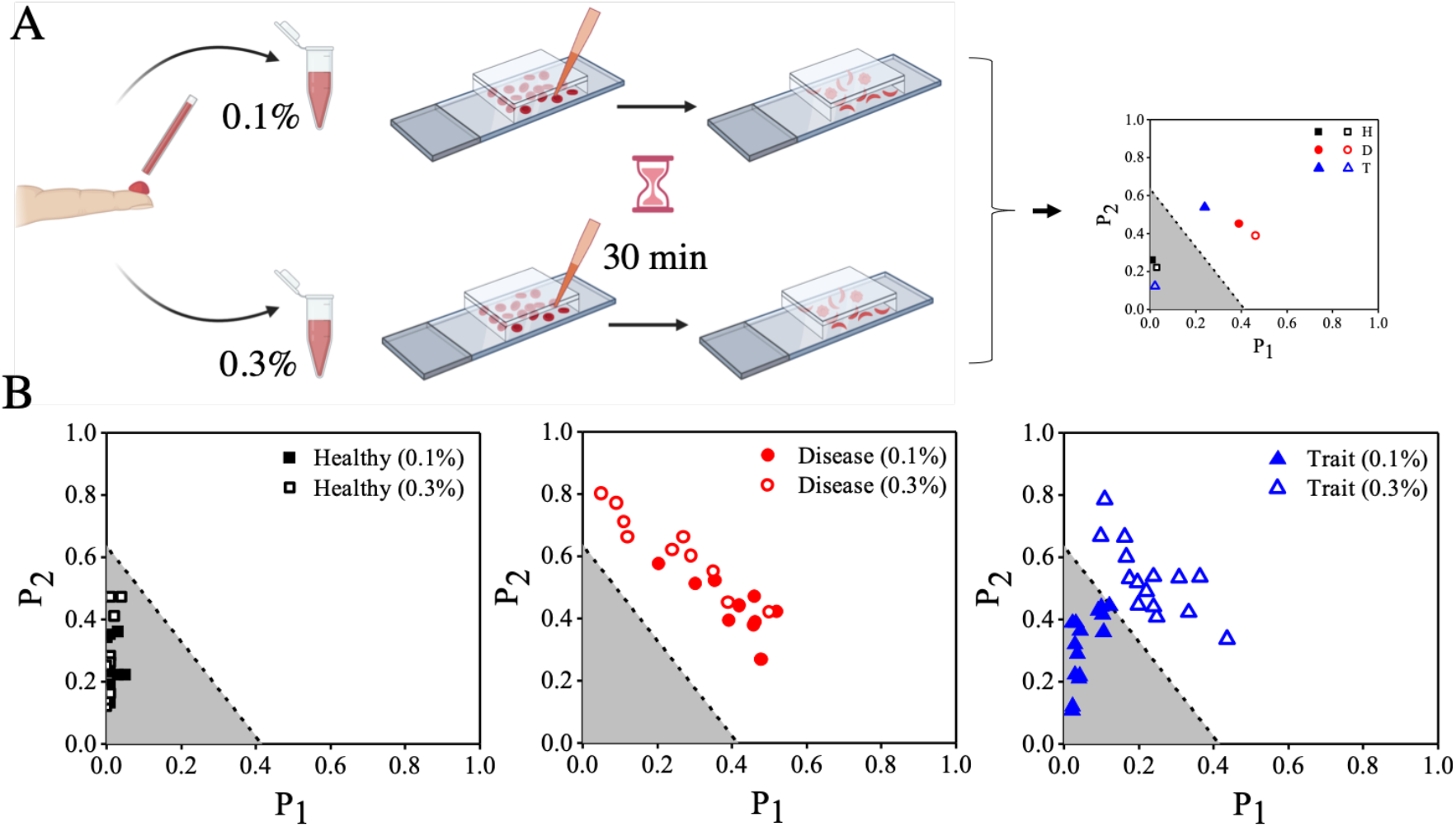
A classification scheme to unambiguously distinguish between disease, trait and healthy blood samples. (A) Workflow to classify an unknown sample. Each unknown sample is treated with two different sodium metabisulphite concentrations (0.1% and 0.3%) and imaged after 30 min. (B) P_2_ vs. P_1_ plots for healthy, disease and trait samples. Solid and hollow symbols correspond to 0.1% and 0.3% sodium metabisulphite respectively. For a healthy sample both points lie below the classifier (grey region). For a disease sample, both points lie above the classifier (white region). For a trait sample, the point corresponding to 0.1% sodium metabisulphite lies below the classifier, while the point corresponding to 0.3% sodium metabisulphite lies above the classifier. (C) Our technique accurately classified 35 distinct unknown samples (H = 10; D = 10 and T = 15) as shown in the three plots.

As shown in **figure 6B**, we validated the classification scheme in a pilot study where we accurately identified thirty-five unknown blood samples (H = 10, D = 10 and T = 15) from adults and older children. We also compared the performance of this classifier with the other classifier (combination #12) identified from the connectivity-Dunn index plot. As shown in **figure S8** in the supporting information, this classifier too could correctly identify all 35 unknown samples. This is the first proof of concept demonstrating RBC shape as the sole biophysical parameter to distinguish between sickle homozygous (disease) and heterozygous (trait) blood samples with high accuracy. However, this technique is yet to be validated with other hemoglobin variants or blood samples obtained from newborns with high levels of fetal hemoglobin.

## Conclusion

We demonstrated a microscopy technique for confirmed diagnosis of sickle cell disease in less than an hour that could be a potential gamechanger in resource-challenged settings by distinguishing between sickle cell disease patients who need immediate clinical intervention (e.g., treatment) and carriers who need non-clinical intervention (e.g., counselling). This technique can change the paradigm of the current two-step diagnosis protocol by exploiting the differential polymerization of hemoglobin in disease and trait blood under controlled hypoxic conditions in a microfluidic chamber and using it to conclusively discriminate between these samples by microscopy alone. Using this technique, we correctly identified 35 blood samples as healthy, sickle cell disease or trait. We built an extremely robust portable smartphone microscope to perform our test in multiple remote field locations. The GPS feature of the smartphone can be utilized for recording the location of new patients. This feature also makes it the only diagnostic technique with built-in disease mapping. Due to faster confirmed results, our test can improve patient compliance to screening efforts and reduce the burden on HPLC systems. The test needs just a drop of blood instead of 2 - 4 ml of venous blood collected for HPLC. While this study was designed for testing blood samples from older children and adults, it is particularly suitable for newborns where only a small volume of blood from a heel-prick is available.

## Data Availability

Suitably de-identified images are available for research purposes from the corresponding author on request.

## Data availability statement

The main data from which conclusions are drawn are included in the manuscript and the supporting information. Suitably de-identified images of blood samples are available for research and teaching purposes from the corresponding author on request.

## Code availability statement

All codes used for analyzing images and processing the data can be found at the following link. https://github.com/ridz46/SickleCellDataAnalysis

## Acknowledgements

The authors would like to thank Sister Lavina, Sister Molly, Sister Monica, Mr. Bhavesh Raicha, Mr. Manoj Parekh, the staff at Shirin and Jamshed Guzder Regional Blood Centre (Valsad), and the staff at Dayanand Hospital (Talasari) for use of leftover blood samples and sharing of de-identified HPLC results. They acknowledge Dr. G. Nageshwara Rao, Dr. Kanjaksha Ghosh, Dr. Roshan Colah, Dr. Malay Mukherjee and Dr. Manisha Madkaikar for technical discussions, Dr. Santosh Noronha for use of dissolved oxygen meter, and Mr. Devendra Dhaka, Mr. Binil Jacob, Mr. Siddhant Jaitpal and Mr. Santosh Jinnawar for other technical and logistical support to the project. This project was supported by a Grand Challenges Explorations (phase 1) grant from Bill and Melinda Gates Foundation through their IKP-GCE program and a translation grant from Tata Centre for Technology and Design (TCTD), Indian Institute of Technology Bombay. Riddha Manna and Anish Mahto received salary support from a grant funded by the Wadhwani Research Centre for Bioengineering (WRCB), Indian Institute of Technology Bombay. The authors are grateful to Dr. Shamik Sen, Dr. Anirban Sain and Dr. Suchita Nath-Sain for their critical feedback on the manuscript. Parts of figures 1, 2 and 6 were created with BioRender.com.

## Author contributions

CD, OS, RM, MS, S, PG and DP designed research. YI provided sickle blood samples and the HPLC results. CD, OS, RM, MS, S, SS, AM, PG, NM, and SS performed research. CD, OS, RM, SS, AM, PG, SS and DP analyzed data. R.M., C.D., O.S., P.G. and D.P. wrote the paper. CD and OS contributed equally to this work.

## Supporting information

### 1. Design and development of an inverted portable smartphone microscope

**Figure S1.**
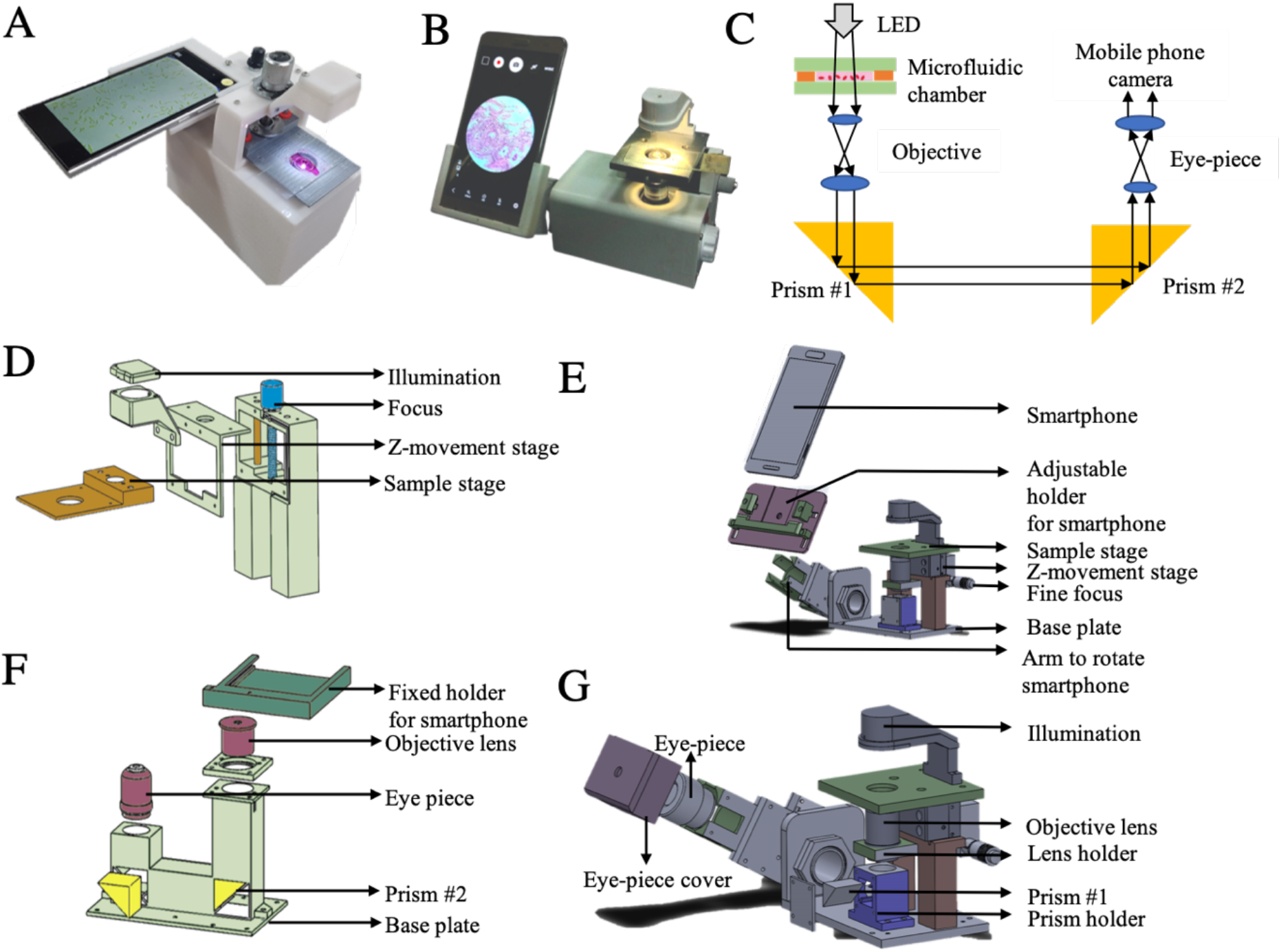
An inverted single-objective portable smartphone microscope. Photos of the microscope models with fixed viewing angle (A) and adjustable viewing angle (B). (C) Ray diagram showing the light path inside both microscopes. (D) and (E) show the exploded 3D schematic of the mechanical parts of (A) and (B) respectively, while (F) and (G) show the corresponding optical systems.

**Figure S1** shows the details of the portable and battery-operated single-objective microscope that we developed. **Figure S1(A)** shows the microscope model where the mobile phone holder is attached to the microscope body at a fixed angle, while the phone holder can be adjusted by any angle from 0° to 90° in the model shown in **Figure S1(B)**. As shown in **figure S1(C)**, a 1W white LED, fitted with a collimating lens, acts as the illumination source. The light transmitted from the microfluidic chip passes through a 40X (0.65 NA) air objective and falls on a right-angled prism (prism #1 in the figure), which bends the beam by 90°. Another right-angled prism (prism #2 in the figure) bends the beam by another 90° such that it falls on an eye-piece lens with 15X magnification. A total magnification of 600X (using a 40X objective and a 15X eye-piece) is achieved in our microscope over a tube length of 160 mm. The smartphone can be removed by the user when the microscope is not in use.

**Figures S1(D)** and **S1(E)** show the exploded views of the mechanical assembly inside (A) and respectively. A microscope slide or a microfluidic chip is placed on the sample stage. The sample stage is attached to a second stage (called “Z-movement stage” in our diagram) capable of vertical movement in both models. This arrangement allows us to move the sample stage up or down by turning the focusing knob. Therefore, the sample stage, and not the objective, is moved while focusing the image in our design. The focusing knob has a pitch of 0.25 mm. The Z-movement stage is fixed to the base plate of the microscope. The entire focusing arrangement and the base plate are made of aluminium to make the microscope stable. The base plate also supports another metallic vertical arm containing the objective lens holder. While the mobile phone holder in (A) is fixed, a key feature of the mechanical system in (B) is the presence of a 3D-printed rotating arm containing the optical components and the smartphone holder. This arm can be rotated by any angle from 0° to 90° to change the viewing angle of the smartphone. The outer casing of the microscope is 3D-printed in PLA.

**Figure S1(F)** and **S1(G)** shows the optical components of (A) and (B) respectively. The system of two prisms makes the microscope design compact. In case of design (B), the rotating arm contains the second prism, the eye-piece and the mobile phone holder. The rotating arm is mounted in such a way that (i) the incident surface of the second prism always remains parallel to the emitting surface of the first prism during rotation, and (ii) the emitting surface of the second prism remains parallel to the eye-piece. The arm rotates in a plane orthogonal to the beam connecting the two prisms. The 3D-printed smartphone holder is detachable and we customized it to the specific mobile phone models. We used a 3D printed PLA cover for the eye-piece to have a fixed distance between the eye-piece and the smartphone camera such that a sharp and focused image forms on the phone screen. The smartphone holder slides onto a slot on the eye-piece cover and keeps the camera in perfect alignment with the rest of the optical system.

### 2. Workflow for image analysis of RBCs

RGB images captured by the smartphone are converted into 8-bit grayscale images. After background subtraction and automatic thresholding, morphological operations are performed to obtain the outlines of the RBCs. Using an area filter, we rule out RBCs with areas <500 pixels (∼2.5 μm^2^) as possible debris > 7500 pixels (∼9.8 μm^2^) as clusters of RBCs. The detailed steps in ImageJ are described below.

The captured image **(1)** is first cropped **(2)** to fit the field of view and converted into an 8-bit grayscale image **(3)**. We then subtract the background with a rolling ball radius of 50 pixels **(4)**. Next, we make the grayscale image into a binary one using the automatic thresholding algorithm of ImageJ. The RBCs appear black on a white background **(5)**. This binary image is them converted into a mask, to make the RBCs appear white against a black background **(6)**.

**Figure S2.**
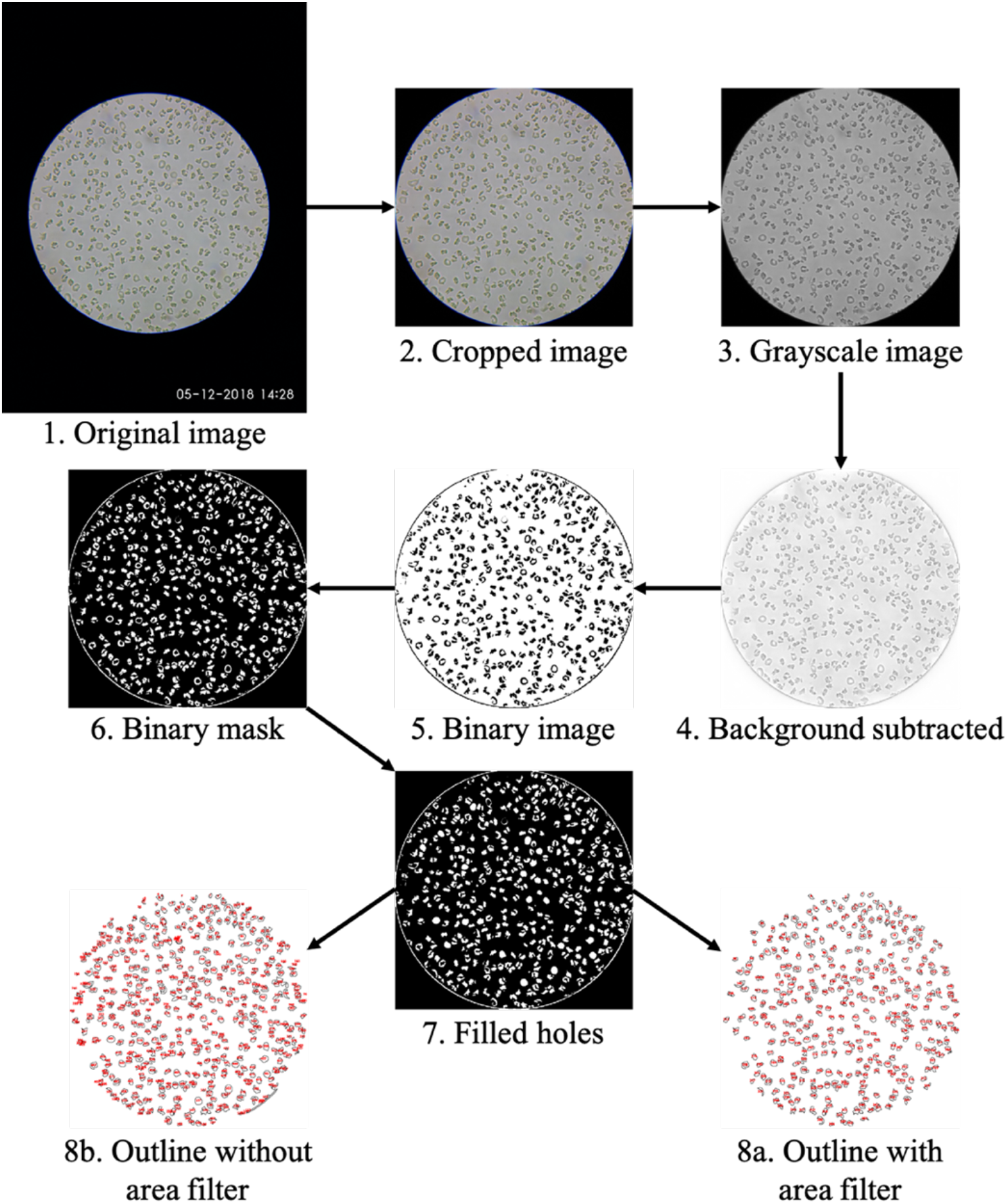
Steps involved in image analysis are shown. The original image (1) is opened in ImageJ and the area of interest is cropped (2). Then image is converted into an 8-bit grayscale image (3) and its background is subtracted to bring the RBCs in the foreground (4). The image is then binarized (5) and converted into a binary mask (6) to get the RBCs as white particles. The holes in the particles are filled to fill the center of the unfocused part of biconcave RBCs (7). Then the RBCs are analyzed to get their solidity, roundness values and outlines. 8a shows the outlines of the RBCs when an area filter is applied to fit the RBC sizes, while 8b shows the outlines without an area filter.

On the masked binary image, we perform the operation ‘fill holes’ **(7)**. This is required as the central part of the biconcave RBCs remain out of focus, making these RBCs appear doughnut-shaped in the binary image. We then analyze the particles (RBCs) to get a list of unique identifiers for each RBC and their shape descriptors including roundness and solidity. Next, we apply an area filter described earlier to exclude debris and connected cells (**8a**). The panel **8b** shows the same cells without the area filter. We then apply a solidity cut-off, where all RBCs with solidity <0.8 are excluded. The RBCs remaining after this step are used to plot the roundness distributions.

### 3. Deoxygenation rate depends on oxygen scavenger concentration

We plotted the dissolved oxygen concentration as a function of time for different sodium metabisulphite concentrations (**figure 3A** in the manuscript). We fitted each plot to an equation of the functional form 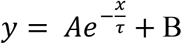, where *y* indicates the dissolved oxygen content (%) and *x=* indicates time (min). **Table S1** shows the fitting parameters for each concentration.

**Table S1:** Fitting parameters to extract the decay time constant (τ) describing the decrease in dissolved oxygen present in RPMI-1640 for different concentrations of the oxygen scavenger sodium metabisulphite.

| *Sodium metabisulphite* | 0.1% | 0.2% | 0.3% | 0.4% | 0.5% |
| --- | --- | --- | --- | --- | --- |
| *A* | 112.5 | 117.1 | 106.6 | 100.7 | 99.6 |
| *B* | - 7.2 | - 4.5 | - 0.9 | 0.2 | 0.8 |
| $\tau$ (min) | 12.2 | 9.3 | 6.1 | 3.5 | 1.6 |

### 4. Choice of diluent and dilution factor

**Figure S3.**
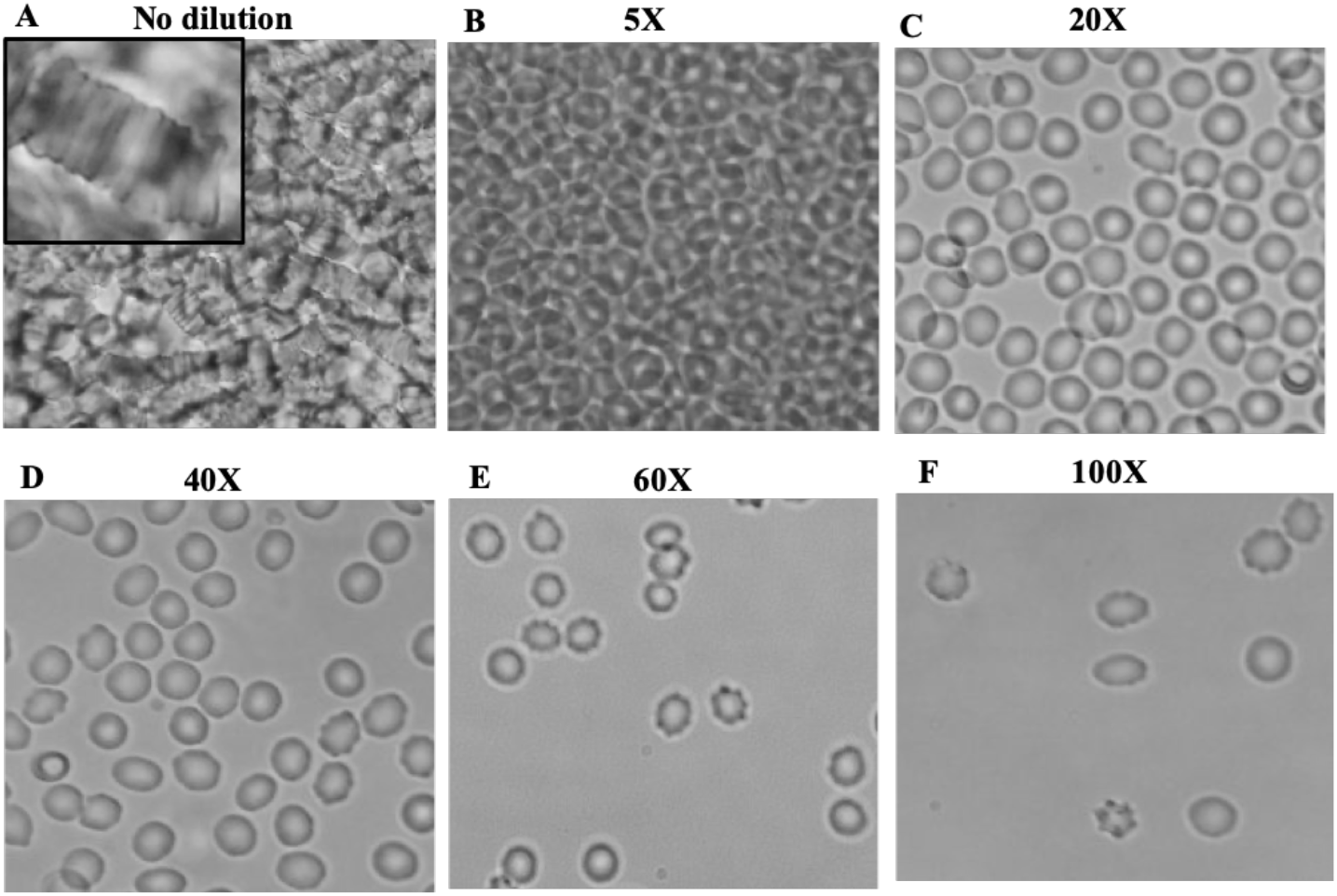
Images of RBCs with different dilutions. (A) Strong rouleaux formation is seen with undiluted blood. (B) There are too many RBCs at 5X dilution, making it difficult for an image processing program to distinguish individual cells. (C) A dilution of 20X gives a sufficient number of cells in the field of view. (D – F) Dilutions of 40X, 60X and 100X lead to very few cells in the field of view, making it difficult to obtain reliable statistical data.

The buffer used to dilute blood must keep RBCs under minimal osmotic stress to avoid crenation, while the dilution factor should ensure that there is an adequate number of RBCs in the field of view. We first diluted healthy RBCs using 0.9% normal saline (NS), 5% dextrose, 1X phosphate buffered saline (PBS) and cell culture media (RPMI-1640). 5% dextrose led to clumping of healthy RBCs and was discarded. Next, sodium metabisulfite solutions of appropriate concentrations as discussed in the manuscript were prepared in NS, PBS and RPMI-1640 and were added to whole blood from trait and disease patients. Compared to RPMI-1640, sickling in PBS and normal saline took 2-3 times longer. Therefore, we continued our study with RPMI-1640 as the diluent.

The dilution was adjusted in such a way that the cells do not form stacks in the imaging chamber. As seen in **figure S3**, whole blood showed strong rouleaux formation. There were too many RBCs present in the field of view with 5X diluted blood, making it difficult for the image processing program to identify individual RBCs. The cells were sparsely distributed for 20X and 40X dilutions. However, there were too few cells in the field of view when blood was diluted by 40X, 60X or 100X. Dilutions higher than 40X also showed an increase in crenation. Therefore, we decided to continue our experiments with 20X dilution.

### 5. Raw images of disease, trait and healthy samples treated with sodium metabisulphite for 30 min

**Figure S4.**
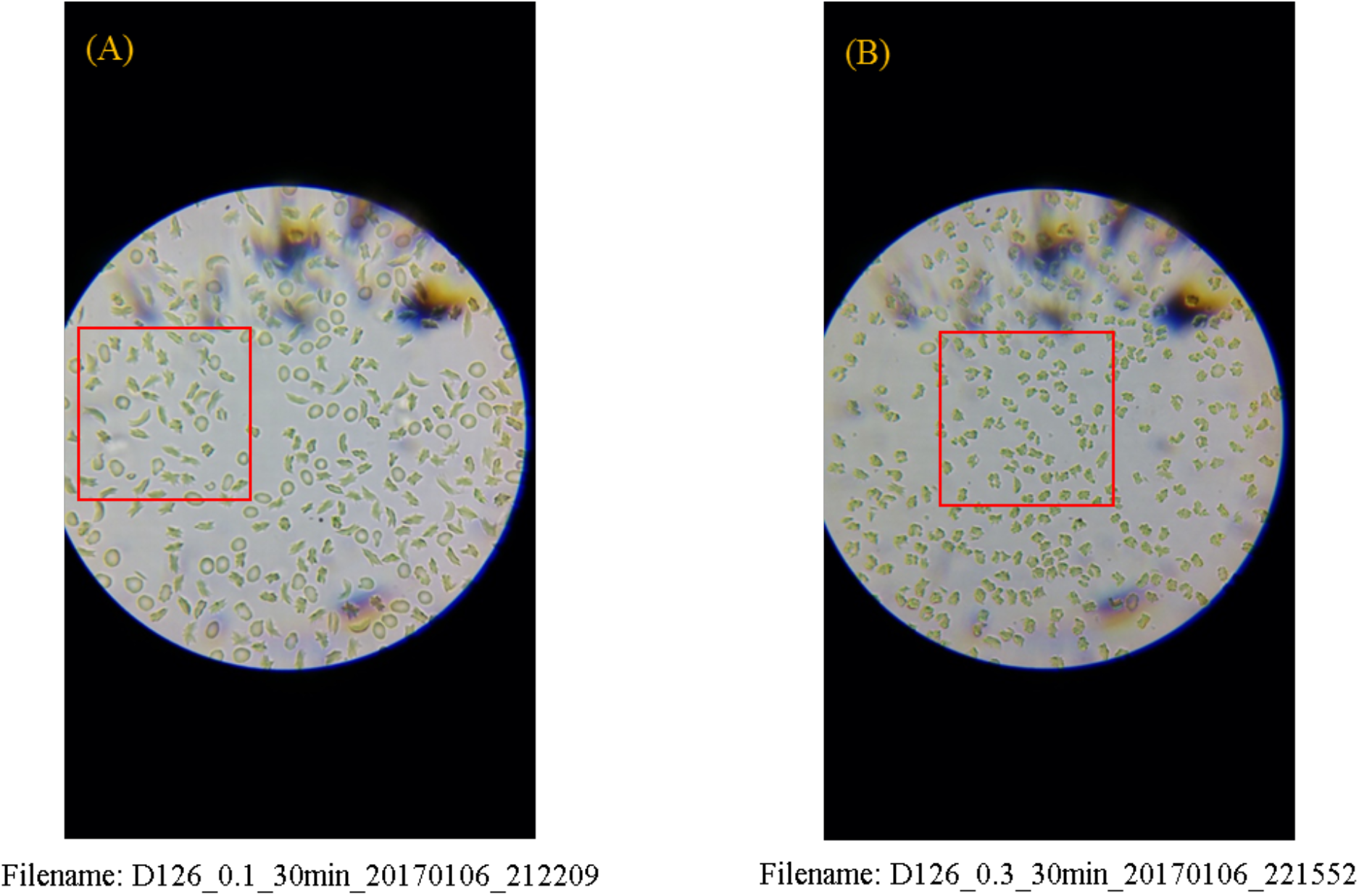
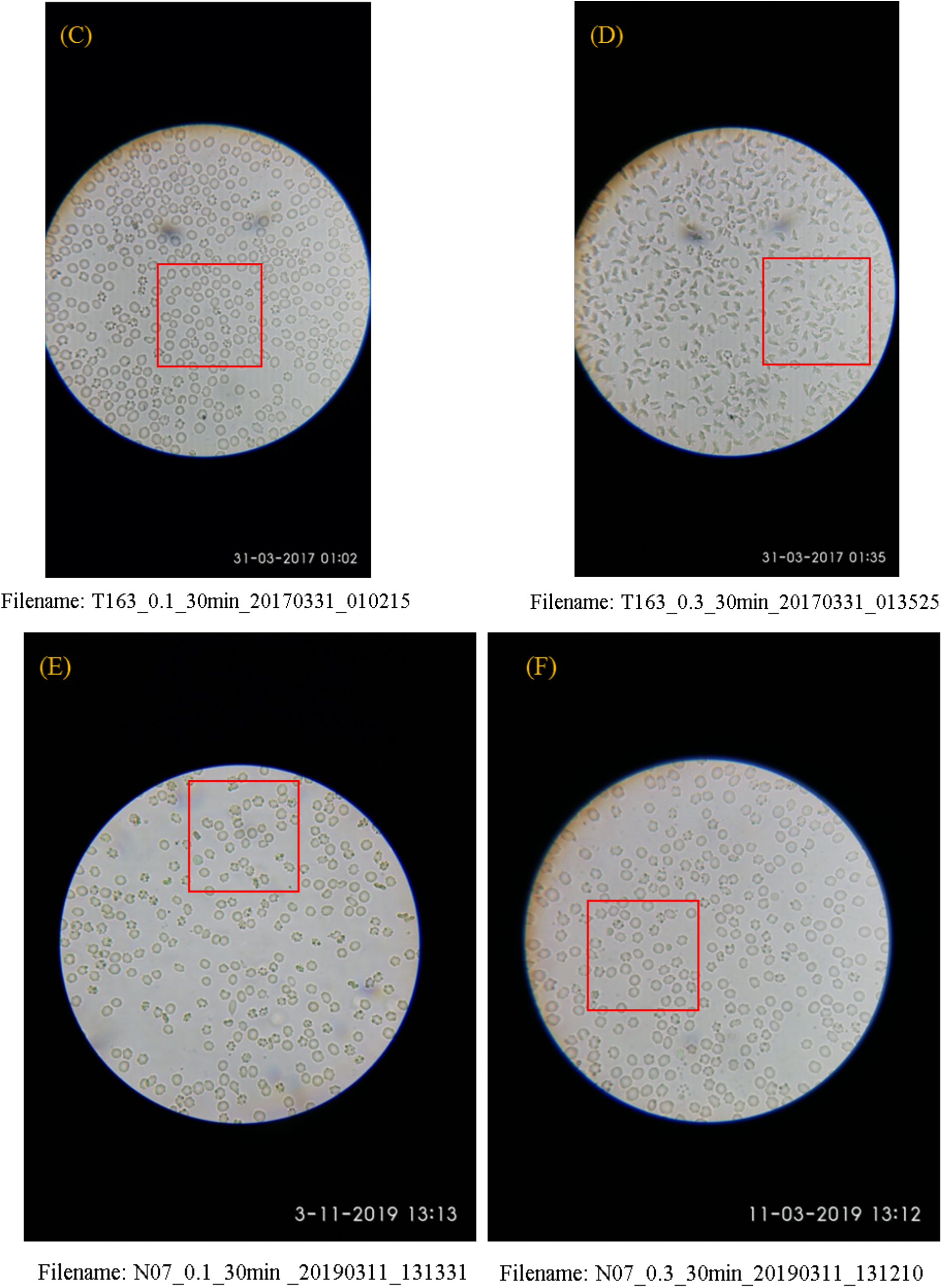
Raw images of disease (A and B), trait (C and D) and healthy (E and F) samples treated with 0.1% (left column) and 0.3% (right column) sodium metabisulphite respectively. The red rectangles show the areas from which snippets were taken for figure 4A.

**Figure S4** shows some representative raw images of disease, trait and healthy blood samples. The red rectangles show the areas that were used as snippets in figure 4 of the manuscript.

### 6. RBC shapes in disease samples treated with 0.1% sodium metabisulphite

When we treat a disease sample with 0.1% sodium metabisulphite, we see holly leaf and granular RBCs in addition to sickle RBCs, as discussed in the manuscript. An image of a representative disease sample was manually annotated by an experienced user in **figure S5**. We find that out of 282 RBCs in the field of view, there are 100 (35%) sickle, 76 (27%) holly leaf and 63 (22%) granular RBCs. There are also 33 unsickled (12%) RBCs and 10 (4%) RBCs that could not be categorized.

**Figure S5.**
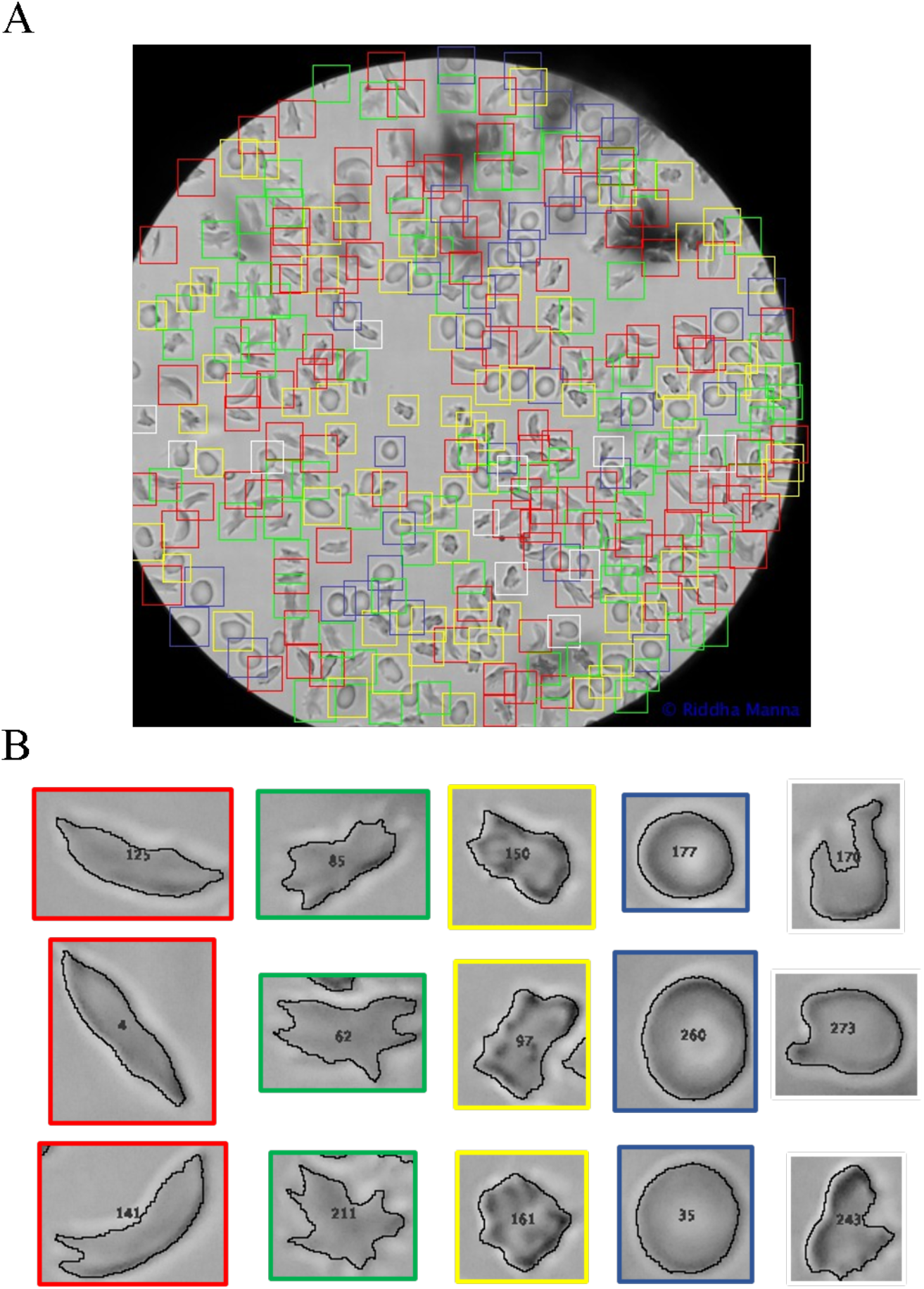
Image of the disease sample (D126) shown in figure S4(A) after manual annotation by an experienced user. (A) We see sickle (red rectangles), holly leaf (green rectangles), granular (yellow rectangles), unsickled (blue rectangles) and uncategorized (white rectangles) RBCs. (B) Three examples of each cell type are shown.

### 7. Choosing roundness (R) over form factor (FF) to characterize RBC shapes

We explored both roundness and form factor for characterising individual RBC shapes in our study. We imaged healthy blood samples and measured both these parameters using ImageJ for RBCs that are very close to circular in shape. Note that the parameter ‘form factor’ (FF) reported by Wheeless and others is the same as the parameter ‘circularity’ (C) measured by ImageJ. Both R and FF range from 0 for elongated objects to 1 for perfect circles.

**Figure S6:**
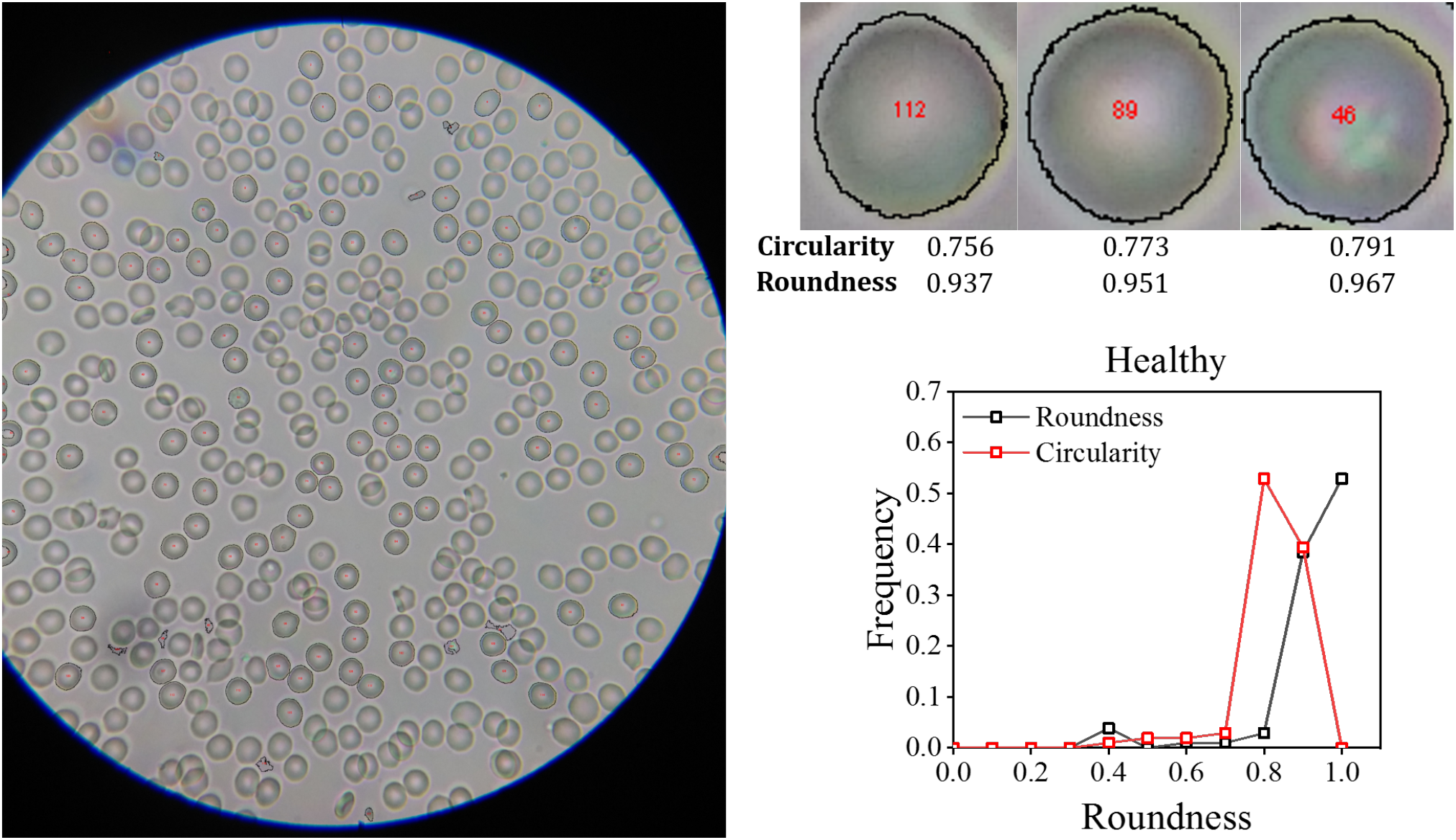
Comparing roundness and form factor for characterizing individual RBC shapes. An image of a healthy flood sample is shown. The circularity and roundness values of three apparently circular RBCs in this image are also shown. As expected, the roundness distribution of the healthy sample peaks at 0.9-1.0, while the form factor (measured as circularity by ImageJ) peaks at 0.8.

**Figure S6** shows the image of a healthy blood sample. Three individual RBCs (numbered 112, 89 and 46) that appear circular in this image are shown separately with their corresponding shape parameters. While the roundness values of these RBCs are >0.9, the form factor is ∼ 0.7. This is because the form factor formula uses the value of the measured perimeter, which is more sensitive to the pixels in the boundary of an object. Therefore, the use of form factor in our image analysis could introduce artifacts in the brightfield images of sickle cells that often have blurred boundaries. On the other hand, the formula for roundness uses the value of the major axis, which is not sensitive to the pixels at the boundary of an object. Therefore, we used roundness to describe the shapes of irregular objects such as sickle RBCs.

### 8. Image artifacts in healthy blood samples leading to tails in roundness distributions

**Figure 4C** of the manuscript shows that the roundness distributions of healthy samples have tails at values of R<0.8. As shown in **figure S7**, this is due to the presence of specific artifacts such as RBCs lying sideways (left panel), crenated RBCs (middle panel) or two or more RBCs overlapping in such a way that the area and solidity filters cannot exclude them (right panel). Our area filter during image processing is chosen to ensure that most sickle cells, which are somewhat larger in area compared to healthy biconcave RBCs, are included in our analysis. This area filter cannot exclude these artifacts when dealing with healthy blood samples. However, the presence of the artifacts does not affect our workflow as seen from the data presented in **figure 5** and **figure 6** of the manuscript.

**Figure S7.**
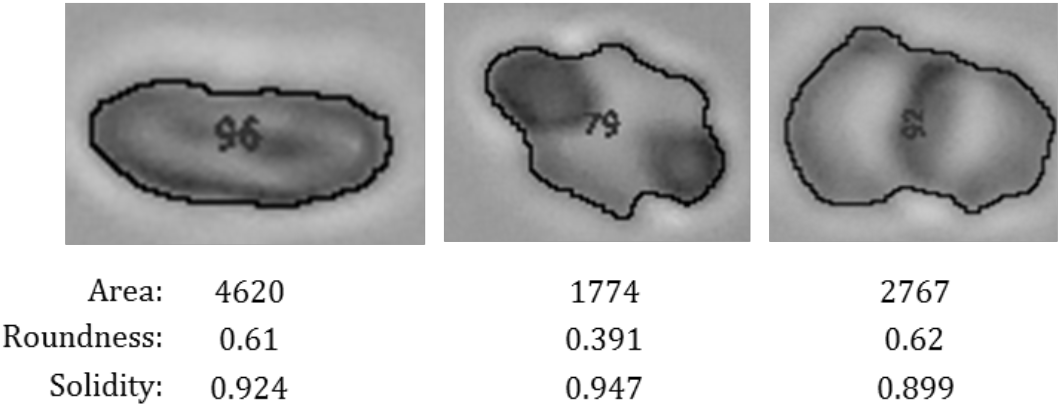
Images of selected RBCs from a healthy sample (N03) showing RBCs lying sideways (left panel), crenated RBCs (middle panel) or overlapping RBCs (right panel).

### 9. *P*_1_ and *P*_2_ combinations used to construct the Dunn index vs. connectivity plot

**Table S2:** Parameters used to construct the Dunn index vs. connectivity plot.

| Combination number | Window size | Roundness range for $P_1$ | Roundness range for $P_2$ | Connectivity | Dunn index |
| --- | --- | --- | --- | --- | --- |
| 1 | 0.2 | 0.0 – 0.2 | 0.2 – 0.4 | 2.01 | 0.07 |
| 2 | 0.2 | 0.1 – 0.3 | 0.3 – 0.5 | 4.97 | 0.02 |
| 3 | 0.2 | 0.2 – 0.4 | 0.4 – 0.6 | 14.75 | 0.03 |
| 4 | 0.2 | 0.3 – 0.5 | 0.5 – 0.7 | 4.56 | 0.07 |
| 5 | 0.2 | 0.4 – 0.6 | 0.6 – 0.8 | 15.07 | 0.08 |
| 6 | 0.2 | 0.5 – 0.7 | 0.7 – 0.9 | 11.31 | 0.07 |
| 7 | 0.2 | 0.6 – 0.8 | 0.8 – 1.0 | 4.03 | 0.06 |
| 8 | 0.3 | 0.0 – 0.3 | 0.3 – 0.6 | 7.07 | 0.02 |
| 9 | 0.3 | 0.1 – 0.4 | 0.4 – 0.7 | 12.42 | 0.05 |
| 10 | 0.3 | 0.2 – 0.5 | 0.5 – 0.8 | 4.16 | 0.08 |
| 11 | 0.3 | 0.3 – 0.6 | 0.6 – 0.9 | 8.15 | 0.06 |
| 12 | 0.3 | 0.4 – 0.7 | 0.7 – 1.0 | 3.45 | 0.14 |
| 13 | 0.4 | 0.0 – 0.4 | 0.4 – 0.8 | 11.01 | 0.09 |
| 14 | 0.4 | 0.1 – 0.5 | 0.5 – 0.9 | 11.36 | 0.02 |
| 15 | 0.4 | 0.2 – 0.6 | 0.6 – 1.0 | 6.77 | 0.02 |
| 16 | 0.5 | 0.0 – 0.5 | 0.5 – 1.0 | 6.30 | 0.00 |

**Table S2** lists the parameters used to plot the Dunn index and connectivity values for sixteen different combinations of *P*_*1*_and *P*_2_. We varied window widths from 0.2 to 0.5. Combination numbers 10 and 12 with window widths of 0.3 each indicate the most promising combinations and these were used to analyse the data of 35 unknown samples.

### 10. An alternate classification scheme

**Figure S8.**
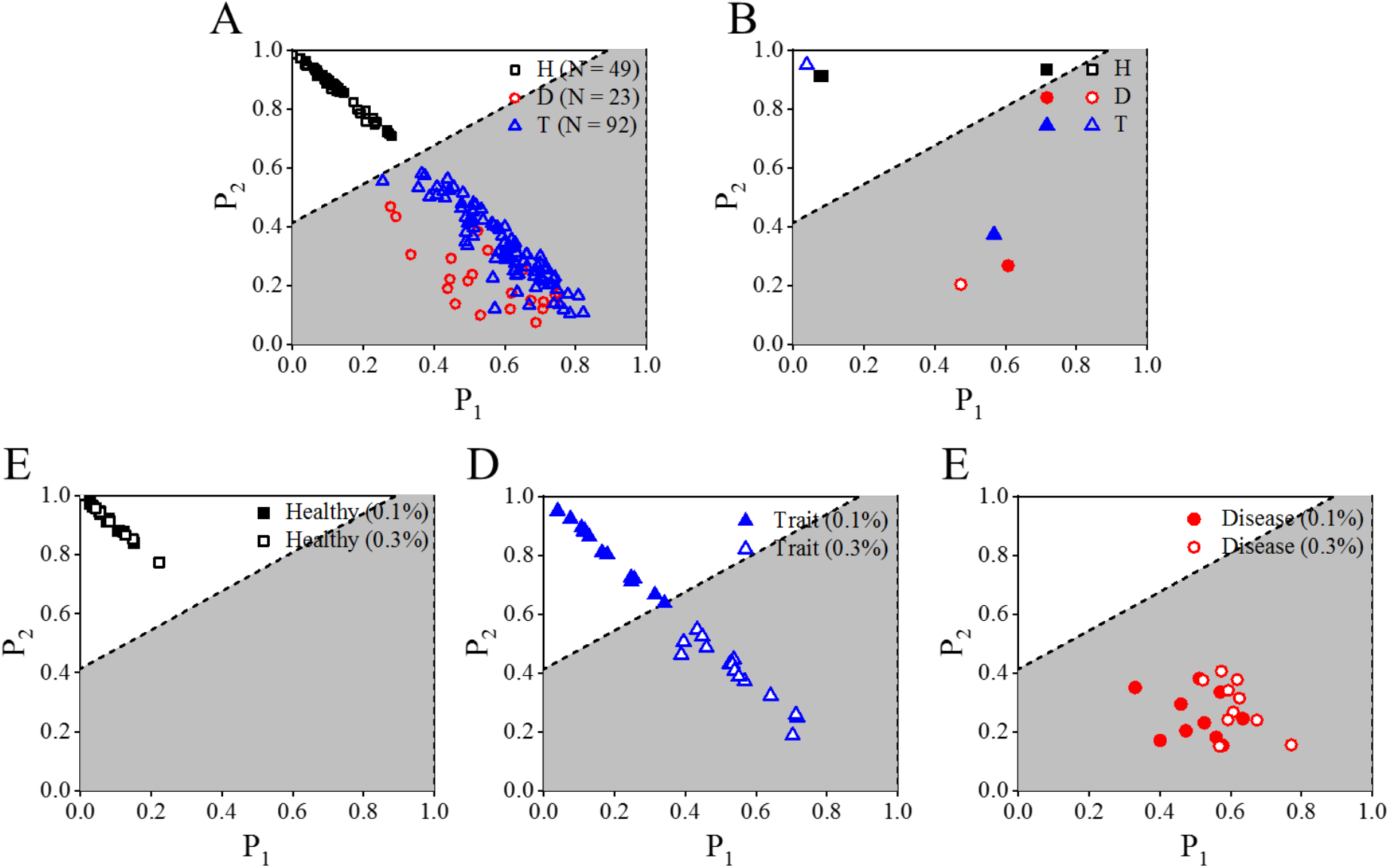
An alternate classifier based on the parameter values of P_**1**_: 0.4 – 0.7 and P_2_: 0.7 – 1.0. (A) In a plot of 164 known samples (H = 49, D = 23 and T = 92), the dotted line indicates the classifier that separates healthy and sickle blood samples. The white area in all the plots indicates the parameter space for healthy samples, while the grey area indicates the parameter space for sickle (disease and trait) samples. (B) Both points corresponding to a healthy sample treated with 0.1% and 0.3% sodium metabisulphite lie above the classifier, while the corresponding points for a disease sample lie below the classifier. The point corresponding to a trait sample treated with 0.1% sodium metabisulphite lies in the space (white area) for healthy samples. The point for a trait sample treated with 0.3% sodium metabisulphite lies in the sickle (grey) zone. (C-E) Validation data for healthy (H = 10), trait (T = 15) and disease (D = 10) samples. All 35 unknown samples were accurately identified by this classifier.

Figure S8 shows an alternate classification scheme given by combination #12 of *P*_*1*_and *P*_2_ values. This classifier also classifies 35 unknown samples with 100% accuracy.

### 12. Captions of movies uploaded as supporting information

**Movie 1:** Sickling video of a disease sample treated with 0.1% sodium metabisulphite. The 30 min video has been sped up 29.5 times. The sample ID is D188.

**Movie 2**: Sickling video of a disease sample treated with 0.3% sodium metabisulphite. The 30 min video has been sped up 29.8 times. The sample ID is D188.

**Movie 3**: Sickling video of a trait sample treated with 0.1% sodium metabisulphite. The 30 min video has been sped up 29.5 times. The sample ID is T187.

**Movie 4**: Sickling video of a trait sample treated with 0.3% sodium metabisulphite. The 30 min video has been sped up 29.5 times. The sample ID is T

## Notes

### Competing Interest Statement

The authors have declared no competing interest.

### Clinical Trial

This was not a clinical trial.

### Author Declarations

All studies using human blood samples were approved by the Institute Ethics Committee (IEC), IIT Bombay, with approval numbers IITB-IEC/2016/016, IITB-IEC/2017/020 and IITB-IEC/2018/042. Written informed consent to use leftover blood samples was taken from all participants.

